# Clinical Performance of Direct RT-PCR Testing of Raw Saliva for Detection of SARS-CoV-2 in Symptomatic and Asymptomatic Individuals

**DOI:** 10.1101/2022.05.15.22275086

**Authors:** Rosa Castillo-Bravo, Noel Lucca, Linyi Lai, Killian Marlborough, Galina Brychkova, Charlie Lonergan, Justin O’Grady, Nabil-Fareed Alikhan, Alexander J. Trotter, Andrew J. Page, Breda Smyth, Peter C. McKeown, Jelena D. M. Feenstra, Camilla Ulekleiv, Oceane Sorel, Manoj Gandhi, Charles Spillane

**Affiliations:** Genetics & Biotechnology Lab, Ryan Institute, National University of Ireland Galway, University Road, Galway H91 REW4, Ireland; Quadram Institute Bioscience, Norwich Research Park, Norwich, Norfolk, UK; College of Medicine, Nursing and Health Sciences, National University of Ireland Galway, Ireland; Health Service Executive (HSE) West, Merlin Park University Hospital, Galway, Ireland; Thermo Fisher Scientific, South San Francisco, USA

## Abstract

RT-qPCR tests based on RNA extraction from nasopharyngeal swab samples are promoted as the “gold standard” for SARS-CoV-2 detection. However, self-collected saliva samples offer a non-invasive alternative more suited to high-throughput testing. This study evaluated the performance of TaqPath COVID-19 Fast PCR Combo Kit 2.0 assay for detection of SARS-CoV-2 in raw saliva relative to a lab-developed direct RT-qPCR test (SalivaDirect-based PCR) and a RT-qPCR test based on RNA extraction from NPS samples. Both samples were collected from symptomatic and asymptomatic individuals (N=615). Saliva samples were tested for SARS-CoV-2 using the TaqPath COVID-19 Fast PCR Combo Kit 2.0 and the SalivaDirect-based PCR, while RNA extracts from NPS samples were tested by RT-qPCR according to the Irish national testing system. The TaqPath™ COVID-19 Fast PCR detected SARS-CoV-2 in 52 saliva samples, of which 51 were also positive with the SalivaDirect-based PCR. 49 samples displayed concordant results with the NPS extraction-based method, while three samples were positive on raw saliva. Among the negative samples, 10 discordant cases were found with the TaqPath COVID-19 Fast PCR (PPA–85.7%; NPA–99.5%), when compared to the RNA extraction-based NPS method, performing similarly to the SalivaDirect-based PCR (PPA-87.5%; NPA-99.5%). The direct RT-qPCR testing of saliva samples shows high concordance with NPS extraction-based method for SARS-CoV-2 detection, providing a cost-effective and highly-scalable system for high-throughput COVID-19 rapid-testing.

## Introduction

The emergence of severe acute respiratory syndrome coronavirus 2 (SARS-CoV-2) in Wuhan in 2019 led to a global pandemic of coronavirus disease 2019 (COVID-19). SARS-CoV-2 can lead to both symptomatic and asymptomatic infections, making detection of infected individuals challenging if based solely on symptomatic diagnostic testing. To combat viral spread and ensure public health, countries have implemented different strategies related to diagnostic, screening and surveillance testing. COVID-19 tests should exhibit high sensitivity and quick turn-around-times to adapt treatment, reduce the spread of disease, and adjust public health interventions to the local epidemiology. Establishing COVID-19 testing in high-throughput settings such as schools or workplaces also requires tests that are easy to use, that require minimal resources and have a high acceptance rate by the individuals involved in the testing^1^.

Detection of SARS-CoV-2 in RNA extracted from nasopharyngeal swab (NPS) samples using quantitative RT-qPCR is considered to be the gold standard for identification of COVID-19 infection, as the virus typically infects the upper respiratory tract. However, reliable collection of NPS requires trained health care professionals, and NPS samples can be difficult to obtain from some individuals due to the discomfort associated with the technique. Using saliva as an alternative sample type to NPS offers several advantages, including non-invasive self-collection, reduced risk of viral transmission and lower sample costs in terms of trained health care personnel, personal protective equipment and costs associated with sample collection^2^.

A number of studies have shown that saliva and NPS RT-PCR-based tests exhibited comparable analytical performance^3-10^. In addition, several reports indicate that saliva might be more sensitive than nasopharyngeal or nasal swabs for diagnosis of SARS-CoV-2 infection, especially for asymptomatic cases or with the emergence of new SARS-CoV-2 variants that can have a different tropism compared to earlier variants^7, 11-13^. Indeed, the 2021 guidance on the use of saliva as sample material for COVID-19 testing highlighted the potential of saliva for nucleic acid based (i.e. PCR based) SARS-CoV-2 testing, while cautioning on the use of saliva as a sample for rapid antigen or antibody tests^14^.

The aim of this retrospective study was to evaluate the performance of the TaqPath™ COVID-19 Fast PCR Combo Kit 2.0 and our SalivaDirect-based (SDB) RT-PCR protocol in raw saliva specimens in comparison to the NPS RNA extraction-based TaqPath™ COVID-19 CE-IVD RT-PCR which is considered to be the gold standard for the detection of SARS-CoV-2.

## Methods

### Clinical specimens

Saliva samples from 615 individuals were collected in the Republic of Ireland (Galway) between February and May 2021 at two locations (Airport Testing Centre and National University of Ireland Galway). All individuals provided a signed informed consent, and the study was approved by the NUI Galway Research Ethics Committee (Approval Number: 2020.08.016; Amend 2102). Of the 615 individuals, 39.7% (N=244) were symptomatic, 35.3% (N=217) were asymptomatic, while the information on the symptomatic status was not available for the remaining part of the cohort (Figure 1A). The Asymptomatic or Symptomatic status of each individual was assigned based on answer given to the question “*Reason why you are being tested by the HSE”* in the registration form that was provided to each volunteers. Each individual who indicated that they had a cough and/or high temperature were classified as symptomatic, while others were classified as asymptomatic (such individuals had been referred for testing as they had been contact traced in accordance with the government guidelines of the time). All saliva samples were tested upon collection using the SalivaDirect-based RT-PCR. Concurrent to saliva collection, NPS were collected and tested for SARS-CoV-2 presence using an RNA extraction-based method according to the national COVID-19 testing system in Ireland run by the Health Service Executive (HSE). Following circa 9 months of storage at -20 °C, raw saliva samples were thawed and re-tested using the lab’s SDB RT-qPCR as well as the TaqPath™ COVID-19 Fast PCR Combo Kit 2.0. Exclusion criteria included: inconclusive result on the TaqPath™ COVID-19 Fast PCR Combo Kit 2.0 and altered status prior to and following storage on the SDB RT-PCR test.

**Figure 1.**
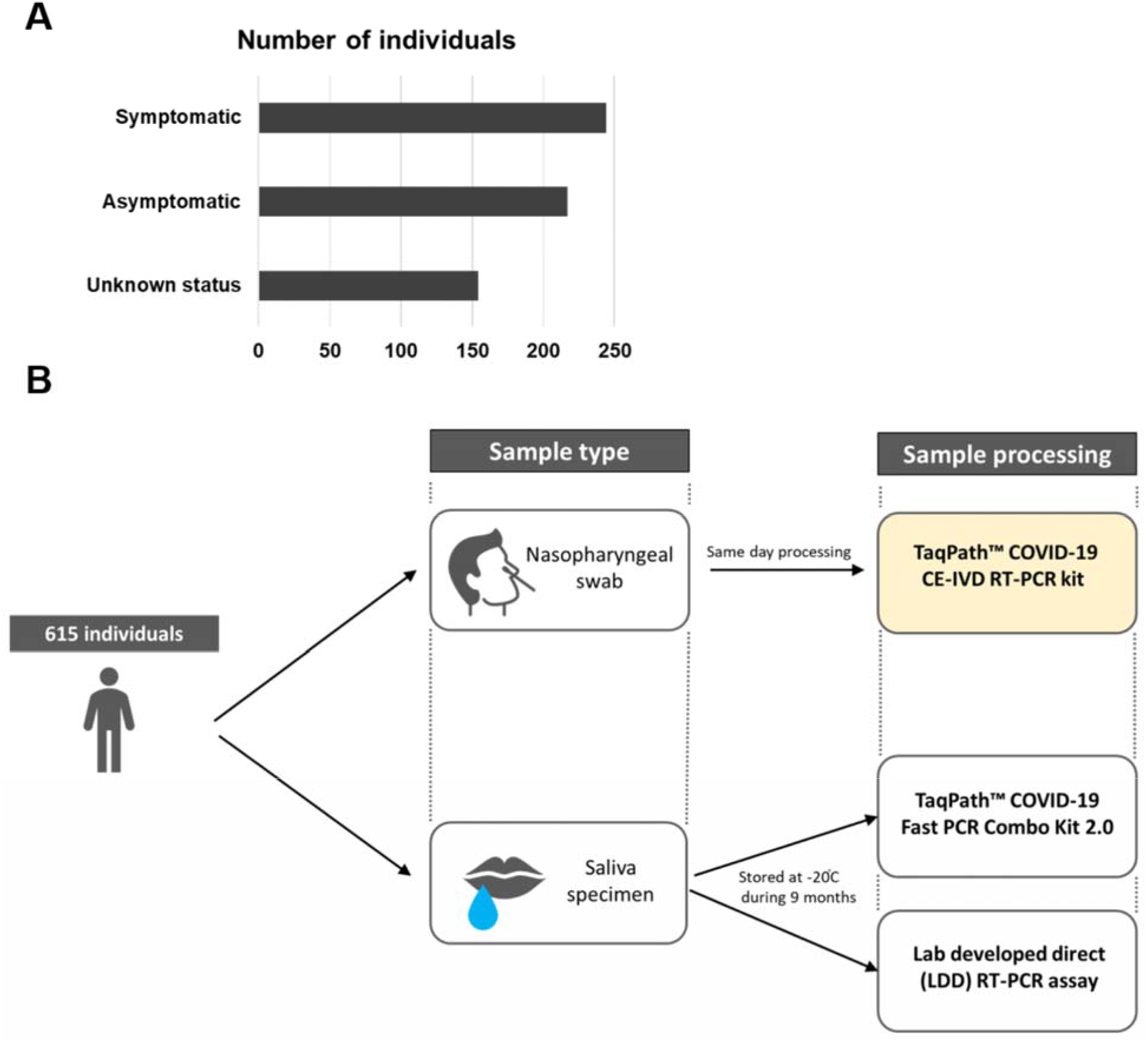
Study design. **A)** Cohort description based on the symptomatic status. **B)** A total of 615 individuals provided saliva and nasopharyngeal swab samples on the same day. Samples were processed according to the algorithm shown.

### SARS-CoV-2 detection

Raw saliva samples were tested upon collection and following storage using the SDB RT-PCR test. In brief, the samples were treated according to the SalivaDirect protocol; 25 uL of each raw saliva sample was collected on a 2.0 mL Eppendorf tube and treated with Proteinase K at 2.5 ug/uL final concentration followed by heat inactivation at 95 °C for 5 minutes. RT-PCR was performed on the Applied Biosystems StepOnePlus™ Real-Time PCR System using the Applied Biosystem TaqMan™ Fast Virus 1-Step Master Mix together with the CDC 2019-Novel Coronavirus Real-time RT-PCR diagnostic panel and results analysed using the StepOne™ Software v2.3. In parallel, saliva samples were tested using the TaqPath™ COVID-19 Fast PCR Combo Kit 2.0 according to the manufacturer’s instructions. The TaqPath™ COVID-19 Fast PCR Combo Kit 2.0 is a fast direct PCR, without RNA extraction, which includes 8 targets across 3 SARS-CoV-2 genes (Orf1a, Orf1b and N) to ensure accurate detection of SARS-CoV-2 as new mutations continue to arise. RT-qPCR was performed on the QuantStudio™ 5 Real Time PCR Instrument with QuantStudio™ Design and Analysis software v1.5.1, and results were analyzed using the Pathogen Interpretive Software CE-IVD Edition 1.1.0. NPS samples were tested within the national HSE testing program using an RNA extraction-based RT-PCR with the TaqPath™ COVID-19 CE-IVD RT-PCR kit. The study design is shown in Figure 1B.

### Whole genome sequencing of SARS-CoV-2

Whole genome sequencing (WGS) of a subset of the SARS-CoV-2 positive saliva samples (N = 46) was performed on RNA extracted from saliva samples via a Quick DNA/RNA Viral MagBead kit (Zymo, R2140). RNA samples were sent on dry ice to the Quadram Institute Bioscience, UK for WGS of SARS-CoV-2. Viral RNA was converted in cDNA and then amplified using the ARTIC protocol v3 (LoCost)^15^ with sequencing libraries prepared using CoronaHiT^16^. WGS was performed using the Illumina NextSeq 500 platform with one positive control and one negative control. The raw reads were demultiplexed using bcl2fastq (v2.20). The reads were used to generate a consensus sequence using the ARTIC bioinformatic pipeline (https://github.com/connor-lab/ncov2019-artic-nf). Briefly, the reads had adapters trimmed with TrimGalore^17^, and were aligned to the Wuhan Hu-1 reference genome (accession MN908947.3) using BWA-MEM (v0.7.17)^18^; the ARTIC amplicons were trimmed and a consensus built using iVAR (v.1.3.0)^19^. Genomes that contained more than 10% missing data were excluded from further analysis to ensure high quality phylogenetic analysis. PANGO lineages were assigned using Pangolin (v2.3.2) (https://github.com/cov-lineages/pangolin) and PangoLEARN model dated 2021-02-21^20^.

## Results

### RT-qPCR on raw saliva shows concordance with RT-qPCR on NPS-extracted RNA for SARS-CoV-2 screening

A total of 615 raw saliva samples obtained from symptomatic and asymptomatic individuals were tested following long-term storage at -20 °C using both the TaqPath™ COVID-19 Fast PCR Combo Kit 2.0 and the lab’s SDB-PCR test. At the time of saliva sample collection, all individuals also provided NPS samples which were tested using an extraction-based RT-qPCR test in an HSE diagnostic laboratory. For all individuals in the study, the result of the RT-qPCR test from the nasopharyngeal swab sample was available. All raw saliva samples were tested both at the time of collection and again following long-term storage at -20 °C using the SDB RT-qPCR assay without an RNA extraction step. For all samples matching results were obtained at the time of sampling and at the time of repeated testing following long-term storage using the SDB RT-qPCR, indicating that no deterioration of sample had occurred.

To evaluate the performance of the direct RT-qPCR testing approach of raw saliva for detection of SARS-CoV-2, results obtained by testing with the TaqPath™ COVID-19 Fast PCR Combo Kit 2.0 were compared to the results based on the nasopharyngeal swab testing using an extraction-based RT-qPCR method (Table 1). SARS-CoV-2 was detected using the TaqPath™ COVID-19 Fast PCR Combo Kit 2.0 in 52 raw saliva samples from the cohort panel, of which 51 were in full agreement with both the SDB-PCR results at the time of collection and re-testing following storage at -20 °C. Interestingly, two samples tested positive only from raw saliva (35<Ct<37). In both cases they tested positive consistently for both the TaqPath™ COVID-19 Fast assay and the SDB-PCR, while the RNA extraction-based testing of the NPS in these cases yielded a negative result.

**Table 1.** Positive and negative percent agreement of the raw saliva-based testing using the TaqPath™ COVID-19 Fast PCR Combo Kit 2.0 and the nasopharyngeal swab-based testing using an RNA-extraction RT-PCR diagnostic assay. Each individual provided one saliva and one nasopharyngeal swab sample on the same day. The nasopharyngeal swabs were processed on the same or following day, while the saliva testing was performed on samples following storage at -20 °C for several months.

|  |  | Extraction based RT-PCR from<br>nasopharyngeal swab samples |  |  |
| --- | --- | --- | --- | --- |
|  |  | Positive | Negative | Total |
| **TaqPath™ COVID-19 Fast**<br>**PCR Combo Kit 2.0** | Positive | 49 | 3 | 52 |
|  | Negative | 10 | 534 | 544 |
|  | Total | 59 | 537 | 596 |
| **Positive Percent Agreement (PPA)** |  | **83.05%** [71.54% to 90.52%] |  |  |
| **Negative Percent Agreement (NPA)** |  | **99.44%** [98.37% to 99.81%] |  |  |

The performance of the lab’s SDB RT-qPCR in raw saliva samples was also evaluated in comparison to the nasopharyngeal swab testing using an extraction-based RT-qPCR method (Table 2) and performed similarly to the TaqPath™ COVID-19 Fast assay.

**Table 2.** Positive and negative percent agreement of the raw saliva-based testing using the SDB RT-PCR assay and the nasopharyngeal swab-based testing using an RNA-extraction RT-PCR diagnostic assay. Each individual provided one saliva and one nasopharyngeal swab sample on the same day. The nasopharyngeal swabs were processed on the same or following day, while the saliva testing was performed on samples following storage at -20 °C for several months.

|  |  | Extraction based RT-PCR from<br>nasopharyngeal swab samples |  |  |
| --- | --- | --- | --- | --- |
|  |  | Positive | Negative | Total |
| **SDB RT-PCR assay** | Positive | 50 | 2 | 52 |
|  | Negative | 9 | 535 | 544 |
|  | Total | 59 | 537 | 596 |
| **Positive Percent Agreement (PPA)** |  | 84.75% [73.48% to 91.76%] |  |  |
| **Negative Percent Agreement (NPA)** |  | 99.63% [98.65% to 99.90%] |  |  |

### Raw saliva-based PCR testing is consistent and can be more sensitive than NPS

SARS-CoV-2 was detected using the TaqPath™ COVID-19 Fast PCR Combo Kit 2.0 in 52 raw saliva samples from the cohort panel, from which 51 were in full agreement with both the SDB-PCR at the time of collection and re-testing following storage at -20□C. Interestingly, 2 samples were positive on raw saliva (35<Ct<37) using both the TaqPath™ COVID-19 Fast assay and the SDB-PCR, while the RNA extraction-based testing of the NPS in these cases showed a negative result.

Whole genome sequencing data was obtained for 46 of the SARS-CoV-2 positive samples. As expected based on the variants circulating in the Republic of Ireland during the period of sample collection (between February 8^th^ and May 6^th,^ 2021), the vast majority of the positive samples consisted of the B.1.1.7 lineage (N=45), with one sample identified as the B.1.562 lineage. When the SARS-CoV-2 clade was determined, 91.1% of the positive samples belonged to the 20I (Alpha, V1) clade, while 8.9% of the samples consisted of the 20A clade. WGS data was available for one of the two samples that showed positivity using both saliva-based testing methods while negative on RNA from NPS, and identified the presence of the Alpha VOC in the sample.

### Saliva-based testing offers good performance for different SARS-CoV-2 detection, including at low viral burden

Of the 52 samples in which SARS-CoV-2 presence was detected using the TaqPath™ COVID-19 Fast PCR Combo Kit 2.0, 42.3% (N=22) of the samples showed a Ct<25, 44.2% (N=23) samples were between 25≤Ct<30 and 13.4% (N=7) of the samples were of low viral load – with Ct≥30 (Figure 2A). Similar sample distribution across the 3 Ct ranges could be observed using the lab’s SDB RT-qPCR (Figure 2A). The comparison of median Ct values in SARS-CoV-2 positive individuals revealed no significant difference between the symptomatic and the asymptomatic patient cohort using both of the RT-qPCR assays used directly on raw saliva samples (Figure 2B-C).

**Figure 2.**
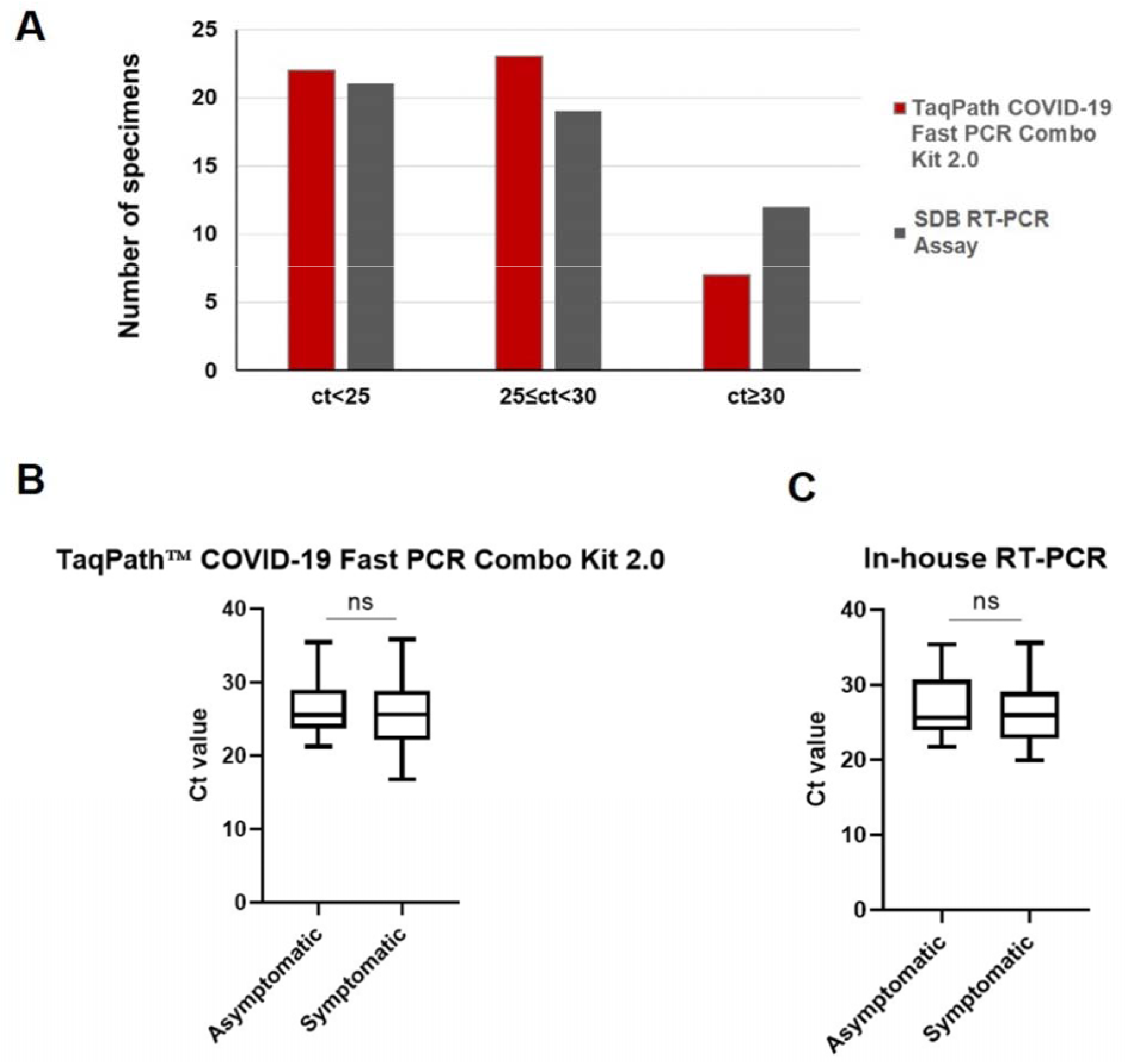
Saliva based SARS-CoV-2 testing Ct values. **A)** Distribution of samples across high, medium and low viral loads grouped by Ct value detected with TaqPath Fast 2.0 kit or SDB RT-PCR. **B-C)** Comparison of the median Ct values between the symptomatic and asymptomatic individuals positive for SARS-CoV-2 using either the TaqPath COVID-19 Fast 2.0 kit **(B)** or the SDB RT-PCR test **(C)**. The box plots show the median (bold horizontal line), interquartile range (box), and total range (whiskers) of detected Ct values.

## Discussion

From the outset of the SARS-CoV-2 pandemic, both nucleic acid and antigen-based tests were developed and deployed, with a major focus on nasopharyngeal swabs as the biological sample of choice to be tested. However, saliva samples are the direct agents of transmission of SARS-CoV-2, through droplets and aerosols, thereby allowing for direct testing for presence of the biological agent within its transmission vehicle. While RT-PCR-based testing of RNA extracted from NPS samples has been considered as the “gold-standard” for SARS-CoV-2 detection, saliva has emerged during the pandemic as a valuable sampling method to improve SARS-CoV-2 detection and workflows^21^. Besides the obvious advantage of self-collection associated with lower costs and reduced risks for viral transmission,^8, 10^ raw saliva samples can be processed directly through RT-qPCR assays which reduces time and removes costs associated with RNA extraction. In addition, saliva (through droplets and aerosols) constitutes a transmission route for SARS-CoV-2 infection and can contain high viral loads of infectious virus as reported by recent studies^7, 11, 12, 22^. Thus, direct testing for SARS-CoV-2 in saliva can help monitor viral loads across variant surges and assess risk of transmission.

Depending on variants, individual factors (genotype, age, health etc) and immunological status (vaccination, prior exposure), SARS-CoV-2 infections can range from asymptomatic to severe symptoms. In the current study we demonstrate that direct RT-qPCR from raw saliva samples using either our in-house developed SDB-PCR assay or a commercially available CE-IVD marked TaqPath Fast kit enables accurate detection of SARS-CoV-2 in both symptomatic and asymptomatic individuals with PPA of >83% and NPA of >99% when compared to the “gold-standard” RNA extraction-based RT-qPCR from nasopharyngeal swabs (Figure 2). Despite the long-term storage (∼ 9 months) of raw saliva samples included in the study, the accuracy between saliva and NPS testing was high. This demonstrates that raw saliva samples can be easily stored for long periods without the need for expensive additives or preservatives.

A number of studies have investigated the use of saliva as a sample method for SARS-CoV-2 detection in comparison to nasopharyngeal swab testing. Although most studies compared RNA extraction-based protocols, the findings of such studies were consistent with our results which used direct RT-qPCR on raw saliva. For saliva samples vs NPS, Pasomsub *et al*. reported a diagnostic sensitivity of 84.3% and specificity of 98.9%^23^, while Yokota *et al*. reported a sensitivity of 92% and specificity of 99.96% for saliva sample versus NPS^24^. Other studies investigating direct PCR saliva based testing obtained comparable results with Moreno-Contreras *et al*. reporting sensitivity of 86.2%^25^ and Vogels *et al*. a positive agreement of 94.1% and a negative agreement of 90.9% for direct PCR from saliva compared to extraction-based NPS testing^26^. Procop *et al*, reported 100% positive agreement (38/38 positive specimens) and 99.4% negative agreement (177/178 negative specimens) by using saliva as specimens from symptomatic patients suspected of having COVID-19^27^. Saliva specimens from Covid-19 confirmed patients even provide greater detection sensitivity and consistency due to an approximately 5X higher viral load compared to nasopharyngeal swabs^10^. Indeed, saliva can offer higher sensitivity and lower variability of saliva testing when compared to the NPS specimens^2, 11^. Our results also demonstrated that two saliva samples gave a positive result using both of the saliva-based direct PCR methods, one of which was confirmed by WGS, while these same samples tested negative on the extraction-based RT-PCR testing of matched nasopharyngeal swabs. This finding suggests that saliva samples may result in greater accuracy from PCR-based testing than nasopharyngeal swabs.

In addition to structural differences between variants at the nucleic or polypeptide levels, the viral load and clearance across tissues and disease stages can potentially differ between variants which in turn could have an impact on what biological specimens are most suitable for detecting different variants. Indeed, the omicron SARS-CoV-2 variant poses a significant challenge for nasal swab based testing as there are indications that saliva based samples may be more effective for diagnostic detection of the omicron SARS-CoV-2 variant relative to NSPs^12, 22^. Marais *et al*. have showed that saliva was a preferred sample for the detection of Omicron variant^11^, which is shown to have an altered tropism for the upper respiratory tract compared to the previous SARS-CoV-2 variants^13^.

The use of a direct-PCR workflows offers an advantage in terms of time-to-result, which in case of both the TaqPath Fast kit and the lab-based SDB assay is under 2 hours. Rapid PCR-based SARS-CoV-2 detection is particularly important in high-frequency testing settings which is often associated with asymptomatic routine testing at workplaces or schools. Our data demonstrate no difference in viral loads between the asymptomatic and symptomatic individuals (Figure 2B-C), in line with previous studies^28^. Several studies have also evaluated the use of RT loop-mediated isothermal amplification (RT-LAMP) in saliva samples for fast detection of SARS-CoV-2. For instance, one study tested different RT-LAMP testing methods using saliva or NPS as sample, and found similar results when using purified/precipitated RNA from each sample type but with significantly reduced sensitivity when the sample is used directly (a reduction from 93% to 65%)^29^. Similar results for direct RT-LAMP were obtained by other groups, indicating significantly lower sensitivity of such approach compared to direct RT-PCR^29-31^.

A desirable feature of any diagnostic kit is the ability to detect different variants, particularly for the case of RNA viruses such as SARS-CoV-2 that are prone to mutation and recombination. While genome sequencing is ideal for characterisation of individual samples, large-scale testing based on genome sequencing has not to date been scaled for everyday practice. All diagnostic tests for SARS-CoV-2 face the challenge of a constantly mutating viral population with periodic emergence of viral variants that display fitness advantages that promote their transmission^32^. For nucleic acid based tests, such challenges to detect new variants arise for homology-based molecular tests (e.g. PCR, LAMP) where the mutations (indels) arise in regions that are detected by sequence homology of the diagnostic test (e.g. the primers)^33^. The emergence of contagious SARS-CoV-2 variants that have undergone significant mutational changes, and display fitness advantages for enhanced transmission in human populations (vaccinated or unvaccinated), can cause surges in COVID-19 cases, as recently exemplified by the Omicron variant. Therefore, there is a high demand for accurate, mutation-resilient, high-throughput testing solutions for both symptomatic and asymptomatic individuals. The development of multiplex assays with several targets across the more conserved regions of the SARS-CoV-2 genome, as for example, the 8 targets in the TaqPath Fast 2.0 assay (targeting Orf1a, Orf1b and N gene) is crucial to enable accurate detection of the virus and avoid false negative testing caused by viral mutations, especially during high surges of cases.

## Conclusions

The current COVID-19 pandemic has highlighted the need for diagnostic testing, screening and surveillance methods that are high-throughput and cost-effective. While point-of-care antigen testing has been deployed at scale globally, the reality is that the detection limit of antigen tests remains poorer than PCR-based methods. However, increasing the throughput of PCR-based testing for more accurate detection of SARS-CoV-2 has been constrained by the use of NPS which are costly and cumbersome to collect. In this study, we demonstrated that highly accurate PCR-based testing can be conducted directly on saliva samples, using a Saliva-Direct based test and a novel CE-IVD marked TaqPath™ COVID-19 Fast PCR Combo Kit 2.0. Saliva-based testing for SARS-CoV-2 provides a highly scalable and accurate approach for rapid detection of SARS-CoV-2 especially during surges of COVID-19 cases, for large-scale mass-testing which includes screening and surveillance programs.

## Data Availability

All data produced in the present work are contained in the manuscript

## Acknowledgements

This study was funded by Science Foundation Ireland grant no. 20/COV/0198 and 21/COV/3753 to CS. We are very grateful to Anne Wyllie (Yale University) and Michael Crone (Imperial College) for support and advice on this work. We are also grateful to the Health Service Executive of Ireland for assistance in sampling.

## References

1. Mina MJ, Andersen KG: COVID-19 testing: One size does not fit all. Science 2021, 371:126–127.

2. Wyllie AL, Fournier J, Casanovas-Massana A, Campbell M, Tokuyama M, Vijayakumar P, Warren JL, Geng B, Muenker MC, Moore AJ: Saliva or nasopharyngeal swab specimens for detection of SARS-CoV-2. New England Journal of Medicine 2020, 383:1283–1286.

3. Bastos ML, Perlman-Arrow S, Menzies D, Campbell JR: The Sensitivity and Costs of Testing for SARS-CoV-2 Infection With Saliva Versus Nasopharyngeal Swabs : A Systematic Review and Meta-analysis. Ann Intern Med 2021, 174:501–510.

4. Butler-Laporte G, Lawandi A, Schiller I, Yao M, Dendukuri N, McDonald EG, Lee TC: Comparison of Saliva and Nasopharyngeal Swab Nucleic Acid Amplification Testing for Detection of SARS-CoV-2: A Systematic Review and Meta-analysis. JAMA Intern Med 2021, 181:353–360.

5. Connor MC, Copeland M, Curran T: Investigation of saliva, tongue swabs and buccal swabs as alternative specimen types to nasopharyngeal swabs for SARS-CoV-2 testing. J Clin Virol 2022, 146:105053.

6. de Paula Eduardo F, Bezinelli LM, de Araujo CAR, Moraes JVV, Birbrair A, Pinho JRR, Hamerschlak N, Al-Hashimi I, Heller D: Self-collected unstimulated saliva, oral swab, and nasopharyngeal swab specimens in the detection of SARS-CoV-2. Clin Oral Investig 2022, 26:1561–1567.

7. Williams E, Bond K, Zhang B, Putland M, Williamson DA: Saliva as a Noninvasive Specimen for Detection of SARS-CoV-2. J Clin Microbiol 2020, 58.

8. Fernandes LL, Pacheco VB, Borges L, Athwal HK, de Paula Eduardo F, Bezinelli L, Correa L, Jimenez M, Dame-Teixeira N, Lombaert IMA, Heller D: Saliva in the Diagnosis of COVID-19: A Review and New Research Directions. J Dent Res 2020, 99:1435–1443.

9. Alqutaibi AY, Saeed MH, Aboalrejal AN: Saliva May Be Considered as Reliable Tool for Diagnosis of COVID-19 When Compared With Nasopharynx or Throat Swabs. J Evid Based Dent Pract 2021, 21:101530.

10. Wyllie AL, Fournier J, Casanovas-Massana A, Campbell M, Tokuyama M, Vijayakumar P, Geng B, Muenker MC, Moore AJ, Vogels CBF, Petrone ME, Ott IM, Lu P, Venkataraman A, Lu-Culligan A, Klein J, Earnest R, Simonov M, Datta R, Handoko R, Naushad N, Sewanan LR, Valdez J, White EB, Lapidus S, Kalinich CC, Jiang X, Kim DJ, Kudo E, Linehan M, Mao T, Moriyama M, Oh JE, Park A, Silva J, Song E, Takahashi T, Taura M, Weizman O-E, Wong P, Yang Y, Bermejo S, Odio C, Omer SB, Dela Cruz CS, Farhadian S, Martinello RA, Iwasaki A, Grubaugh ND, Ko AI: Saliva is more sensitive for SARS-CoV-2 detection in COVID-19 patients than nasopharyngeal swabs. medRxiv 2020:2020.2004.2016.20067835.

11. Marais Gert, Hsiao Nei-yuan, Iranzadeh Arash, Doolabh Deelan, Enoch Annabel, Chun-yat Chu, Williamson Carolyn, Brink Adrian, Diana H: Saliva swabs are the preferred sample for Omicron detection. medRxiv 2021.

12. Adamson B, Sikka R, Wyllie AL, Premsrirut P: Discordant SARS-CoV-2 PCR and Rapid Antigen Test Results When Infectious: A December 2021 Occupational Case Series. medRxiv 2022.

13. Hui KPY, Ho JCW, Cheung MC, Ng KC, Ching RHH, Lai KL, Kam TT, Gu H, Sit KY, Hsin MKY, Au TWK, Poon LLM, Peiris M, Nicholls JM, Chan MCW: SARS-CoV-2 Omicron variant replication in human bronchus and lung ex vivo. Nature 2022.

14. Control ECfDPa: Considerations for the use of saliva as sample material for COVID-19 testing. Stockholm: ECDC, 2021.

15. Quick J: NCoV-2019 Sequencing Protocol v3 (LoCost). Protocolsio 2020.

16. Baker DJ, Aydin A, Le-Viet T, Kay GL, Rudder S, de Oliveira Martins L, Tedim AP, Kolyva A, Diaz M, Alikhan NF, Meadows L, Bell A, Gutierrez AV, Trotter AJ, Thomson NM, Gilroy R, Griffith L, Adriaenssens EM, Stanley R, Charles IG, Elumogo N, Wain J, Prakash R, Meader E, Mather AE, Webber MA, Dervisevic S, Page AJ, O’Grady J: CoronaHiT: high-throughput sequencing of SARS-CoV-2 genomes. Genome Med 2021, 13:21.

17. Krueger F: 2020. FelixKrueger/TrimGalore. Perl. 2016.

18. Heng L: Aligning Sequence Reads, Clone Sequences and Assembly Contigs with BWA-MEM. ArXiv 2013:1303.3997 [q-Bio].

19. Grubaugh ND, Gangavarapu K, Quick J, Matteson NL, De Jesus JG, Main BJ, Tan AL, Paul LM, Brackney DE, Grewal S, Gurfield N, Van Rompay KKA, Isern S, Michael SF, Coffey LL, Loman NJ, Andersen KG: An amplicon-based sequencing framework for accurately measuring intrahost virus diversity using PrimalSeq and iVar. Genome Biol 2019, 20:8.

20. Rambaut A, Holmes EC, O’Toole Á, Hill V, McCrone JT, Ruis C, du Plessis L, Pybus OG: A dynamic nomenclature proposal for SARS-CoV-2 lineages to assist genomic epidemiology. Nat Microbiol 2020, 5:1403–1407.

21. Tan SH, Allicock O, Armstrong-Hough M, Wyllie AL: Saliva as a gold-standard sample for SARS-CoV-2 detection. Lancet Respir Med 2021, 9:562–564.

22. Lai J, German J, Hong F, Tai S-HS, McPhaul KM, Milton DK, Group ftUoMSR: Comparison of Saliva and Mid-Turbinate Swabs for Detection of COVID-19. medRxiv 2022:2021.2012.2001.21267147.

23. Pasomsub E, Watcharananan SP, Boonyawat K, Janchompoo P, Wongtabtim G, Suksuwan W, Sungkanuparph S, Phuphuakrat A: Saliva sample as a non-invasive specimen for the diagnosis of coronavirus disease 2019: a cross-sectional study. Clin Microbiol Infect 2021, 27:285.e281-285.e284.

24. Yokota I, Shane PY, Okada K, Unoki Y, Yang Y, Inao T, Sakamaki K, Iwasaki S, Hayasaka K, Sugita J, Nishida M, Fujisawa S, Teshima T: Mass Screening of Asymptomatic Persons for Severe Acute Respiratory Syndrome Coronavirus 2 Using Saliva. Clin Infect Dis 2021, 73:e559–e565.

25. Moreno-Contreras J, Espinoza MA, Sandoval-Jaime C, Cantú-Cuevas MA, Barón-Olivares H, Ortiz-Orozco OD, Muñoz-Rangel AV, Hernández-de la Cruz M, Eroza-Osorio CM, Arias CF, López S: Saliva Sampling and Its Direct Lysis, an Excellent Option To Increase the Number of SARS-CoV-2 Diagnostic Tests in Settings with Supply Shortages. J Clin Microbiol 2020, 58.

26. Vogels CBF, Watkins AE, Harden CA, Brackney DE, Shafer J, Wang J, Caraballo C, Kalinich CC, Ott IM, Fauver JR, Kudo E, Lu P, Venkataraman A, Tokuyama M, Moore AJ, Muenker MC, Casanovas-Massana A, Fournier J, Bermejo S, Campbell M, Datta R, Nelson A, Yale IRT, Dela Cruz CS, Ko AI, Iwasaki A, Krumholz HM, Matheus JD, Hui P, Liu C, Farhadian SF, Sikka R, Wyllie AL, Grubaugh ND: SalivaDirect: A simplified and flexible platform to enhance SARS-CoV-2 testing capacity. Med (N Y) 2021, 2:263–280 e266.

27. Procop GW, Shrestha NK, Vogel S, Van Sickle K, Harrington S, Rhoads DD, Rubin BP, Terpeluk P: A Direct Comparison of Enhanced Saliva to Nasopharyngeal Swab for the Detection of SARS-CoV-2 in Symptomatic Patients. J Clin Microbiol 2020, 58.

28. Zuin M, Gentili V, Cervellati C, Rizzo R, Zuliani G: Viral Load Difference between Symptomatic and Asymptomatic COVID-19 Patients: Systematic Review and Meta-Analysis. Infect Dis Rep 2021, 13:645–653.

29. Uribe-Alvarez C, Lam Q, Baldwin DA, Chernoff J: Low saliva pH can yield false positives results in simple RT-LAMP-based SARS-CoV-2 diagnostic tests. PLoS One 2021, 16:e0250202.

30. Nagura-Ikeda M, Imai K, Tabata S, Miyoshi K, Murahara N, Mizuno T, Horiuchi M, Kato K, Imoto Y, Iwata M, Mimura S, Ito T, Tamura K, Kato Y: Clinical Evaluation of Self-Collected Saliva by Quantitative Reverse Transcription-PCR (RT-qPCR), Direct RT-qPCR, Reverse Transcription-Loop-Mediated Isothermal Amplification, and a Rapid Antigen Test To Diagnose COVID-19. J Clin Microbiol 2020, 58:e01438–01420.

31. Taki K, Yokota I, Fukumoto T, Iwasaki S, Fujisawa S, Takahashi M, Negishi S, Hayasaka K, Sato K, Oguri S, Nishida M, Sugita J, Konno S, Saito T, Teshima T: SARS-CoV-2 detection by fluorescence loop-mediated isothermal amplification with and without RNA extraction. J Infect Chemother 2021, 27:410–412.

32. Markov PV, Katzourakis A, Stilianakis NI: Antigenic evolution will lead to new SARS-CoV-2 variants with unpredictable severity. Nat Rev Microbiol 2022:1–2.

33. Rajib SA, Ogi Y, Hossain MB, Ikeda T, Tanaka E, Kawaguchi T, Satou Y: A SARS-CoV-2 Delta variant containing mutation in the probe binding region used for RT-qPCR test in Japan exhibited atypical PCR amplification and might induce false negative result. J Infect Chemother 2022, 28:669–677.

